# Manitoba Interdisciplinary Lactation Center (MILC): A bench-to-population human milk biorepository and research platform protocol

**DOI:** 10.64898/2026.02.27.26347256

**Authors:** Larisa C Lotoski, Spencer R Ames, Alie Johnston, Kelsey Fehr, Meghan B Azad

## Abstract

**Introduction:** Breastfeeding supports multiple aspects of child development and maternal health. However, research findings are often inconsistent due to methodological limitations, including inadequate control for sociodemographic factors, variation in feeding practices, health conditions across the life course, and heterogeneity in human milk (HM) composition. The Manitoba Interdisciplinary Lactation Center (MILC) is a globally accessible, bench–to–population research platform that enables integrated study of HM composition, maternal-child health, and the societal and structural determinants of lactation and HM feeding.

**Methods and Analysis:** MILC combines cross-sectional questionnaire data and HM sample collection with longitudinal administrative data derived from provincial government databases. MILC recruits lactating parents currently feeding their HM to at least one child. Participants follow a standardized full breast expression protocol. All collected HM samples have their macronutrient profiles characterized and are bio-banked for unspecified future research. Questionnaires capture child and parent demographic, dietary and health characteristics, and detailed HM feeding practices. Administrative data include over 90 databases spanning health and social services utilization and education; these de-identified records are housed at the Manitoba Population Research Data Repository and linked with MILC study samples and data. MILC questionnaires and HM collection protocols can be customized to accommodate specific research projects (e.g. additional surveys or questions; snap freezing, addition of preservatives, cell or extra-cellular vesicle isolation, etc.). MILC began recruiting participants in October 2024 and is currently ongoing. Researchers may access MILC data and biospecimens subject to appropriate ethical approvals and data–sharing agreements.

**Ethics and dissemination:** MILC is approved by the University of Manitoba Human Research Ethics Board and the Provincial Health Research Privacy Committee. Participation is voluntary and based on informed consent. Research updates and findings will be disseminated via peer-reviewed journal publications, academic and clinical conferences, social media, public knowledge sharing events (e.g. information booths and virtual “Ask Me Anything” sessions), the MILC website (https://www.milcresearch.com) and the MILC Club (monthly meetings among researchers, trainees, healthcare providers, and community partners). MILC members also engage with agenda-setting organizations (e.g. Breastfeeding Committee for Canada, North American Board for Breastfeeding and Lactation Medicine) to accelerate translation of research knowledge into policy and practice.

**STRENGTHS AND LIMITATIONS OF THIS STUDY:**

- MILC combines low-burden cross-sectional human milk samples and questionnaire data with lifelong/longitudinal administrative data.
- Parent-child dyad human milk feeding practices and history are captured in a high level of detail, filling a gap frequently experienced in human milk and lactation research.
- Our questionnaires have been partially harmonized with other biorepositories and/or utilize valid and reliable measurement scales.
- The initial MILC study pilot population lacks diversity; this will be intentionally addressed going forward.
- The cost to maintain a long-term biorepository facility is high.

## INTRODUCTION

Human milk (HM) supplies optimal infant nutrition and breastfeeding is associated with a reduced risk for many prevalent and costly chronic diseases, including asthma and obesity (in breastfed children), as well as type 2 diabetes and breast cancer (in mothers who breastfed their children) (1). Based on this evidence, the World Health Organization recommends exclusive breastfeeding the first 6 months of life and continued breastfeeding for up to 2 years or beyond (2). However, more than 70% of mother-infant dyads in Canada do not meet these recommendations (3), which has substantial health and economic consequences at the population level. In the US, where breastfeeding rates are similar to Canada, suboptimal breastfeeding costs the healthcare system an estimated $3 billion (USD) per year in direct medical costs (4,5). Thus, it is critically important to understand (i) how HM supports child health and development, and more broadly, (ii) the barriers and facilitators that influence breastfeeding uptake and continuation.

While health outcomes associated with breastfeeding are well-documented, the underlying social and biological mechanisms underlying these relationships are not fully understood and considerable heterogeneity exists within breastfeeding research (6–8). This uncertainty has been attributed to methodological flaws, including: i) failure to account for confounding variables (e.g. sociodemographic factors, maternal health); ii) inaccurate health outcome measures (e.g. self- or parent-reported); iii) selection bias due to high rates of attrition in longitudinal studies; iv) retrospective data collection; and v) lack of detailed feeding practice data. Moreover, existing studies rarely distinguish between direct breastfeeding (at the breast) and expressed breastfeeding (from a bottle), and rarely specify the type of complementary feeding for ‘partially breastfed’ infants (i.e. formula vs. solid foods) – yet research by our team and others (9–13) show these nuances are relevant in breastfeeding research and may account for heterogeneity in research results. Also, epidemiologic studies rarely have access to HM samples to interrogate the biological mechanisms underlying observed associations. All of these limitations will be addressed by the Manitoba Interdisciplinary Lactation Centre (MILC).

MILC is a globally unique “bench to population” facility that links high-resolution demographic characteristic data and HM samples to lifelong provincial administrative data in Manitoba, Canada **(Figure 1)**. MILC participants complete questionnaires capturing detailed infant feeding practices, sociodemographic, health and lifestyle information prospectively. HM samples are collected to enable mechanistic studies. Additionally, questionnaire and HM composition data are linked to a wealth of administrative health and social data housed at the Manitoba Population Research Data Repository. The Repository comprises over 90 databases spanning health, social services, criminal justice, and education, and can be linked at an individual-level across these domains and through time (14,15). MILC is accessible to researchers worldwide, providing an unprecedented platform and facility to address critical questions in HM science and maternal and child health.

**Figure 1.**
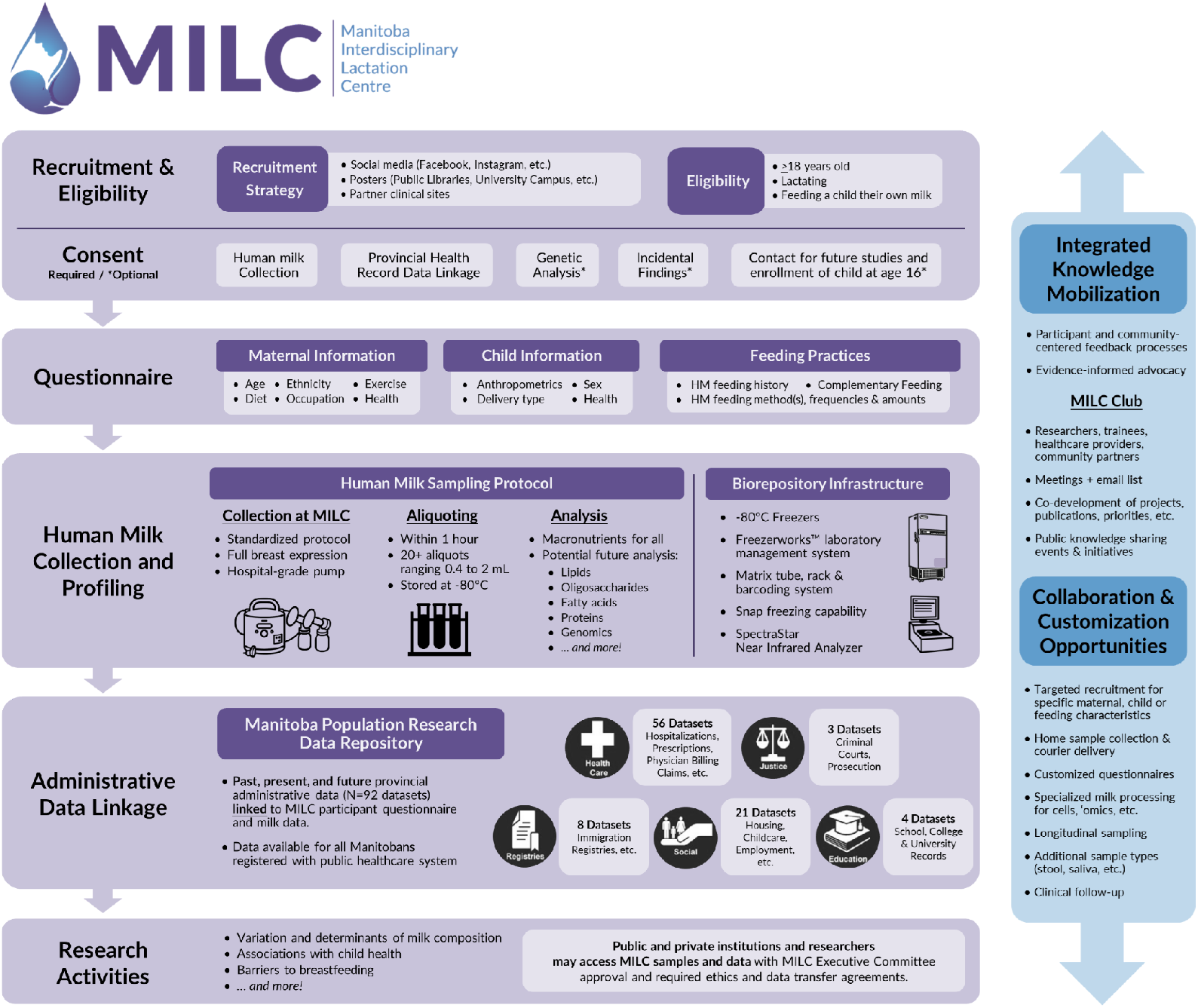
Overview of Manitoba Interdisciplinary Lactation Centre (MILC) workflow and capabilities. HM, human milk.

## METHODS & ANALYSIS

### Study Design & Setting

MILC is a biorepository and research platform situated at the University of Manitoba Bannatyne Medical Campus in Winnipeg, Manitoba, Canada. MILC combines cross-sectional questionnaires and HM sample collection (at any period during lactation) with lifelong retrospective and prospective longitudinal administrative data housed at the Manitoba Population Research Data Repository.

### Recruitment, Consent, Participation & Long-Term Follow-up

Social media marketing (Facebook, Instagram), physical posters (in public libraries, university campuses, etc.), and partner clinical sites are used to recruit general Manitoba population participants. Targeted recruitment for specific maternal, child, or feeding characteristics may be conducted depending on potential research projects. All recruitment methods provide a direct link to the MILC website’s study sign-up form (https://www.milcresearch.com/donate-milk.html). Following confirmation of eligibility (mother is at least 18 years old, lactating, and feeding a child their own HM), digital written informed consent for the mother-child dyad is provided by the participating mother and captured using REDCap (Research Electronic Data Capture) (16,17). Consenting mothers provide their contact information, complete questionnaires about themselves and their participating child (**Table 1; Supplementary Table 1**), provide a HM sample, and permit linkage of provincial administrative data for themselves and the participating child, all with the understanding that their samples and data will be part of unspecified future research and accessible to other researchers, institutions and private/commercial bodies. Optionally, participating mothers may also consent to (i) genetic analyses, (ii) reporting of incidental findings, (iii) receiving study updates and newsletters, (iv) being contacted about future studies, and (v) having their child contacted when they reach the age of 16 years so they can provide their own consent for continued involvement in the study. MILC began recruiting participants in October 2024 and is currently ongoing.

**Table 1.** MILC questionnaire descriptions.

| **Enrollment Questionnaire** |  |
| --- | --- |
| **Consent** | *Required:* Questionnaire and milk collection; Provincial health record data linkage<br>*Optional:* Genetic analysis; Incidental findings; Permission to be contacted for future study participation; Contact child at age 16 for independent consent. |
| **Maternal Demographics** | Age; gender; height; weight; ethnic origins; marital status; years living in Canada. |
| **Maternal Health** | Pregnancy information (past, present); parity; medical conditions (e.g. chronic diseases, allergies); vaccinations; family medical history. |
| **Socioeconomic Status** | Employment status; household income; education level. |
| **Child Health** | Medical conditions (e.g. serious illnesses or infections, birth defects); vaccinations. |
| **Child Demographics** | Age; biological sex; birth weight; gestational age; delivery type (e.g. vaginal, c-section). |
| **Day of Milk Collection Questionnaire** |  |
| **Maternal Health** | Health and symptoms in the last two weeks. |
| **Maternal Medications** | Current medication type, dosage, schedule, and consumption route. |
| **Maternal Physical Activity & Sleep** | Time spent sitting, walking, and doing vigorous or moderate* activity per week; hours of sleep per day. |
| **Maternal Exposures** | Diet (rapid eating assessment survey**, common allergen consumption 24h recall); alcohol, cannabis, nicotine. |
| **Child Health** | Health and symptoms in the last two weeks. |
| **Human Milk Collection** | Collection time; time since feeding or pumping; expression details (full or partial; left or right breast); milk amount collected (mL); expression method (pump or hand). |
| **Feeding** | Human milk feeding method and frequency; complementary feeding. |
| **Maternal Breastfeeding Information** | Current breastfeeding status; previous breastfeeding experience; breastfeeding duration; sources of breastfeeding support; breastfeeding goals. |

\*Vigorous = Heavy lifting, digging, aerobics; Moderate = Carrying light loads, aerobics; \*\*Gans et al. 2003.

### Data Collection: MILC Questionnaires

Mother participants complete two questionnaires. The first questionnaire is deployed immediately following consent and captures mother and child demographic characteristics (maternal characteristics: ethnic origins, immigrant status, marital status, employment and/or student status, household income before and during parental leave, anthropometric measures, past pregnancies, gender; participating child characteristics: birth characteristics, anthropometric measures, medical conditions, diagnoses, illnesses or infections; **Table 1, Supplementary Table 1**). The second questionnaire is completed by mothers on the same day as their HM sampling appointment and captures time-sensitive information (maternal characteristics: diet, current medical conditions and symptoms (in the last 14 days), substance and medication use, physical and sleep behavior, last HM expression, tandem feeding, feeding supports and knowledge sources; child characteristics: current medical conditions and symptoms, historical and current HM feeding practices). Maternal physical activity and sleep behaviours are captured using the International Physical Activity Questionnaire Short Form (18). Maternal diet in the last 14 days is captured using the Rapid Eating Assessment (19). Mothers are also asked to report the ingestion of 13 common allergenic foods within 24 hours of HM sampling, as well as their ingestion or use of alcohol, cannabis, cigarettes and nicotine products, recreational drugs, and prescribed, behind or over-the-counter medications and supplements. Child feeding practices, including the consumption of HM, commercial infant formula, cow’s milk, non-dairy milk, and solid foods, are captured ‘over the last seven days. HM feeding is differentiated by method (directly from the breast/chest vs. pumped or expressed and fed with a bottle or other container). (**Table 1, Supplementary Table 1)**. Questionnaires can be customized to accommodate future research projects and collaborator requests.

### Human Milk Samples: Collection, Processing, Compositional Profiling & Long-Term Storage

Participants typically attend the MILC facility to donate their HM sample. With guidance from our MILC Study Coordinator, HM is collected using a standardized full breast expression protocol using a double electric pump [Symphony® PLUS Breast pump w/Initiation Program Card, Medela, Mississauga, Canada] and sterile pump kits (Medela, 24-mm breast shield). When requested, participants are provided with alternate breast shield sizes (18, 21, 27, 30 mm PersonalFit™ PLUS breast shield, Medela, Mississauga, Canada). The breast is cleaned with sanitary wipes prior to expression. HM is collected from the fullest breast (i.e. the breast not used for the most recent feeding) under low light conditions. A full breast expression is collected. If less than 25 mL are collected, the second breast is cleaned, fully expressed, and HM from both breasts is mixed together. The start time of collection is recorded. Sample collection bottles are wrapped in aluminum foil to reduce sample light exposure.

HM samples are stored in a cooler, on ice, and immediately transported down the hall to the biorepository facility on ice in a light-proof insulated cooler, where they are stored at 4°C until aliquotted (typically within 1 hour of collection; processing time is recorded). At least 20 aliquots (ranging from 0.4-2mL each) are prepared under minimal light to further reduce sample light exposure. Whole HM aliquots are stored in a -80°C Thermo Scientific TSX Series -80°C ultra-low temperature freezer (Thermo Fisher Scientific, Waltham, MA, USA). All HM samples entering the MILC biorepository have their macronutrient profiles characterized by NIR spectroscopy (SpectraStar XT, KPM Analytics, Westborough, USA) in batches after a single freeze/thaw cycle.

HM sampling and analysis protocols can be adjusted to accommodate prospective biobanking and subsequent analyses or assays that require additional pre-processing measures (e.g. snap freezing, skimming, addition of preservatives, cell or extra-cellular vesicle isolation, fractionation, etc.). MILC has the ability to collect additional sample types (e.g. stool, saliva, etc.) depending on future research projects or collaborator requests.

### Administrative Data Linkage

Administrative data is housed at the Manitoba Population Research Data Repository situated at the Manitoba Center for Health Policy (MCHP; Winnipeg, Canada), which holds a collection of over 90 databases covering education (e.g. enrollment records, University and College student data, etc.), health (e.g. immunization, healthcare utilization, comorbidities, mental health diagnoses, etc.), justice (e.g. corrections offender management systems, criminal court networks, etc.), registries (e.g. immigration, health insurance, Indigenous registries, etc.), social (housing assists, disability services, etc.), and support file (urban environment data, long term care facility data, etc.) domains (**Supplementary Table 2**; (14,15,20)). This longitudinal administrative data is primarily derived from Manitoba Government departments, including Manitoba Health, Manitoba Families, Manitoba Education & Training, and Manitoba Justice. The Repository contains information on nearly all Manitoba residents extending as far back as 1970 (21,22). MILC Study participants provide their Provincial Health Information Number (PHIN) and permission to link their study data with their administrative data at MCHP. De-identified MILC data are shared with MCHP for linkage (via PHIN) and analysis within the secure MCHP environment.

### Data Storage & Access

Identifiable participant consent and questionnaire data are permanently stored on the University of Manitoba’s REDCap secure questionnaire server. De-identified consent and questionnaire data are stored in the THRiVE Discovery Lab Database (THRiVEdb, https://www.thrivedb.ca/, Amazon Simple Storage Service (S3), hosted by Amazon Web Services) (23). HM sample aliquot data are managed using Freezerworks laboratory information management system (Dataworks Development Inc., Mountlake Terrace, USA).

Public and private institutions and researchers may access MILC samples and/or questionnaire data with MILC Executive Committee and institutional research ethics board approval, as well as completed material and data transfer agreements. Access to provincial administrative data requires further approval by the Manitoba Provincial Health Research Privacy Committee and institutional research ethics boards, as well as material and data transfer agreements with the MCHP.

MILC encourages prospective collaborations, including those that require sample and data collection beyond current MILC protocols. Where possible, data generated from HM samples are submitted to MILC to facilitate future data sharing. Access inquiries can be directed to.

### Analysis: Pilot Population

One hundred mother-child dyads were recruited to MILC between October 2024 and March 2025 (**Tables 2 and 3**) as a pilot study population. Among optional consents, incidental findings and permission to be contacted for future study participation had the highest consent rates (99.0% and 98.0%, respectively), while receiving bi-annual newsletters (87.0%) had the lowest.

**Table 2.**
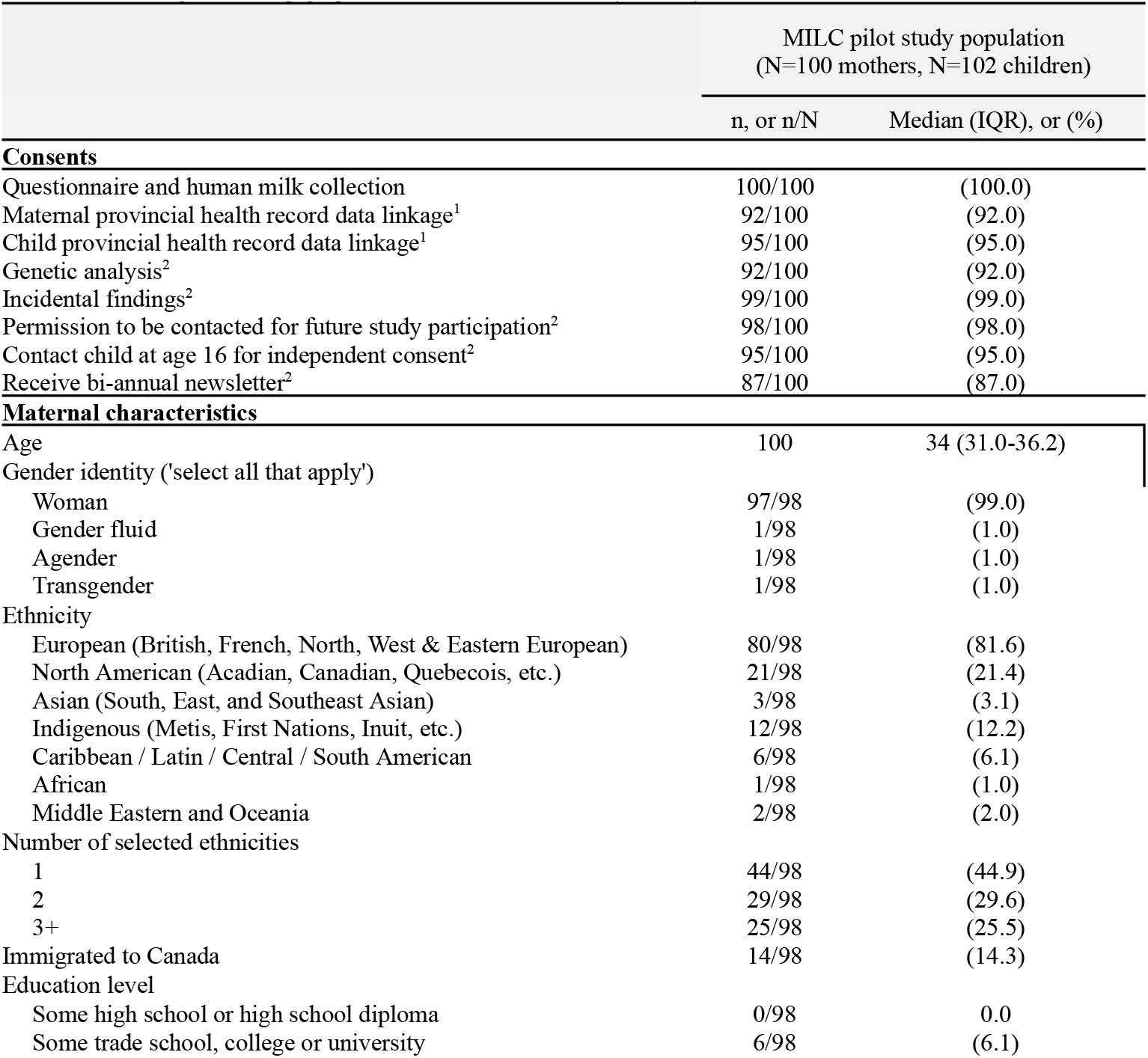
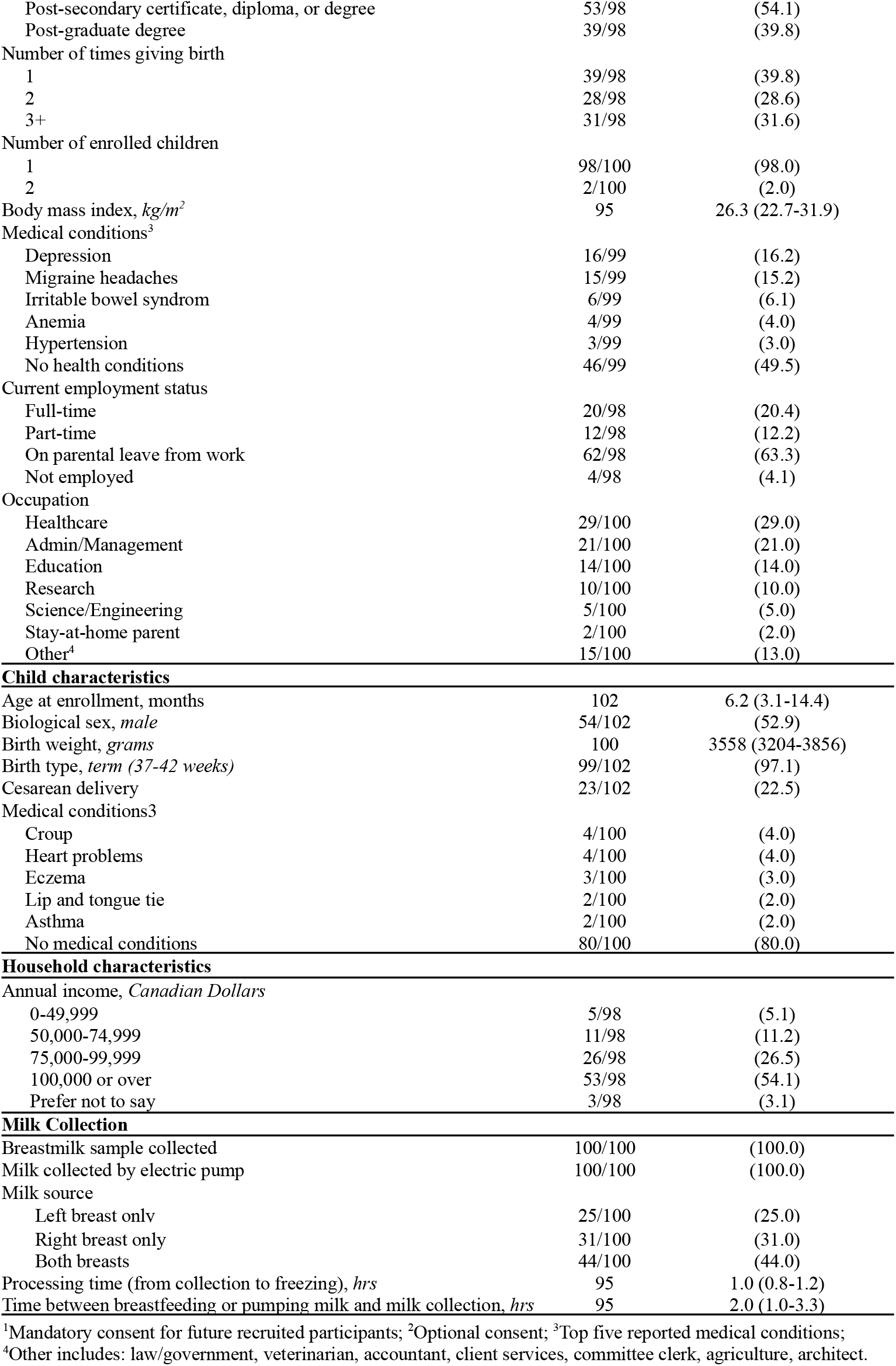
MILC pilot study population characteristics (N=100).

|  | MILC pilot study population<br>(N=100 mothers, N=102 children) |  |
| --- | --- | --- |
|  | n, or n/N | Median (IQR), or (%) |
| **Consents** |  |  |
| Questionnaire and human milk collection | 100/100 | (100.0) |
| Maternal provincial health record data linkage<sup>1</sup> | 92/100 | (92.0) |
| Child provincial health record data linkage<sup>1</sup> | 95/100 | (95.0) |
| Genetic analysis<sup>2</sup> | 92/100 | (92.0) |
| Incidental findings<sup>2</sup> | 99/100 | (99.0) |
| Permission to be contacted for future study participation<sup>2</sup> | 98/100 | (98.0) |
| Contact child at age 16 for independent consent<sup>2</sup> | 95/100 | (95.0) |
| Receive bi-annual newsletter<sup>2</sup> | 87/100 | (87.0) |
| **Maternal characteristics** |  |  |
| Age | 100 | 34 (31.0-36.2) |
| Gender identity ('select all that apply') |  |  |
| Woman | 97/98 | (99.0) |
| Gender fluid | 1/98 | (1.0) |
| Agender | 1/98 | (1.0) |
| Transgender | 1/98 | (1.0) |
| Ethnicity |  |  |
| European (British, French, North, West & Eastern European) | 80/98 | (81.6) |
| North American (Acadian, Canadian, Quebecois, etc.) | 21/98 | (21.4) |
| Asian (South, East, and Southeast Asian) | 3/98 | (3.1) |
| Indigenous (Metis, First Nations, Inuit, etc.) | 12/98 | (12.2) |
| Caribbean / Latin / Central / South American | 6/98 | (6.1) |
| African | 1/98 | (1.0) |
| Middle Eastern and Oceania | 2/98 | (2.0) |
| Number of selected ethnicities |  |  |
| 1 | 44/98 | (44.9) |
| 2 | 29/98 | (29.6) |
| 3+ | 25/98 | (25.5) |
| Immigrated to Canada | 14/98 | (14.3) |
| Education level |  |  |
| Some high school or high school diploma | 0/98 | 0.0 |
| Some trade school, college or university | 6/98 | (6.1) |
| Post-secondary certificate, diploma, or degree | 53/98 | (54.1) |
| Post-graduate degree | 39/98 | (39.8) |
| Number of times giving birth |  |  |
| 1 | 39/98 | (39.8) |
| 2 | 28/98 | (28.6) |
| 3+ | 31/98 | (31.6) |
| Number of enrolled children |  |  |
| 1 | 98/100 | (98.0) |
| 2 | 2/100 | (2.0) |
| Body mass index, $kg/m^2$ | 95 | 26.3 (22.7-31.9) |
| Medical conditions<sup>3</sup> |  |  |
| Depression | 16/99 | (16.2) |
| Migraine headaches | 15/99 | (15.2) |
| Irritable bowel syndrom | 6/99 | (6.1) |
| Anemia | 4/99 | (4.0) |
| Hypertension | 3/99 | (3.0) |
| No health conditions | 46/99 | (49.5) |
| Current employment status |  |  |
| Full-time | 20/98 | (20.4) |
| Part-time | 12/98 | (12.2) |
| On parental leave from work | 62/98 | (63.3) |
| Not employed | 4/98 | (4.1) |
| Occupation |  |  |
| Healthcare | 29/100 | (29.0) |
| Admin/Management | 21/100 | (21.0) |
| Education | 14/100 | (14.0) |
| Research | 10/100 | (10.0) |
| Science/Engineering | 5/100 | (5.0) |
| Stay-at-home parent | 2/100 | (2.0) |
| Other<sup>4</sup> | 15/100 | (13.0) |
| **Child characteristics** |  |  |
| Age at enrollment, months | 102 | 6.2 (3.1-14.4) |
| Biological sex, *male* | 54/102 | (52.9) |
| Birth weight, *grams* | 100 | 3558 (3204-3856) |
| Birth type, *term (37-42 weeks)* | 99/102 | (97.1) |
| Cesarean delivery | 23/102 | (22.5) |
| Medical conditions<sup>3</sup> |  |  |
| Croup | 4/100 | (4.0) |
| Heart problems | 4/100 | (4.0) |
| Eczema | 3/100 | (3.0) |
| Lip and tongue tie | 2/100 | (2.0) |
| Asthma | 2/100 | (2.0) |
| No medical conditions | 80/100 | (80.0) |
| **Household characteristics** |  |  |
| Annual income, *Canadian Dollars* |  |  |
| 0-49,999 | 5/98 | (5.1) |
| 50,000-74,999 | 11/98 | (11.2) |
| 75,000-99,999 | 26/98 | (26.5) |
| 100,000 or over | 53/98 | (54.1) |
| Prefer not to say | 3/98 | (3.1) |
| **Milk Collection** |  |  |
| Breastmilk sample collected | 100/100 | (100.0) |
| Milk collected by electric pump | 100/100 | (100.0) |
| Milk source |  |  |
| Left breast only | 25/100 | (25.0) |
| Right breast only | 31/100 | (31.0) |
| Both breasts | 44/100 | (44.0) |
| Processing time (from collection to freezing), *hrs* | 95 | 1.0 (0.8-1.2) |
| Time between breastfeeding or pumping milk and milk collection, *hrs* | 95 | 2.0 (1.0-3.3) |

<sup>1</sup>Mandatory consent for future recruited participants; <sup>2</sup>Optional consent; <sup>3</sup>Top five reported medical conditions;
<sup>4</sup>Other includes: law/government, veterinarian, accountant, client services, committee clerk, agriculture, architect.

**Table 3.**
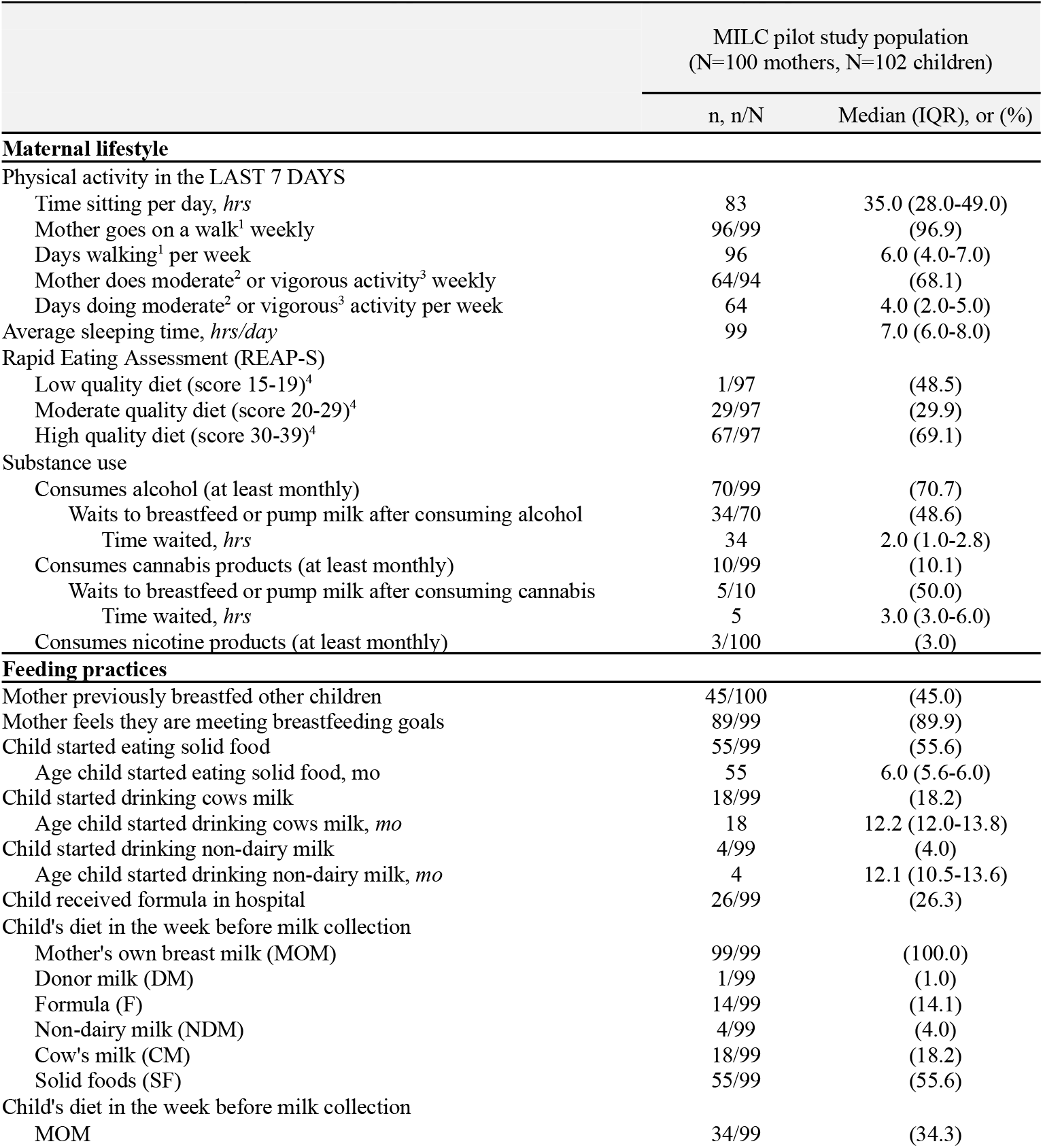
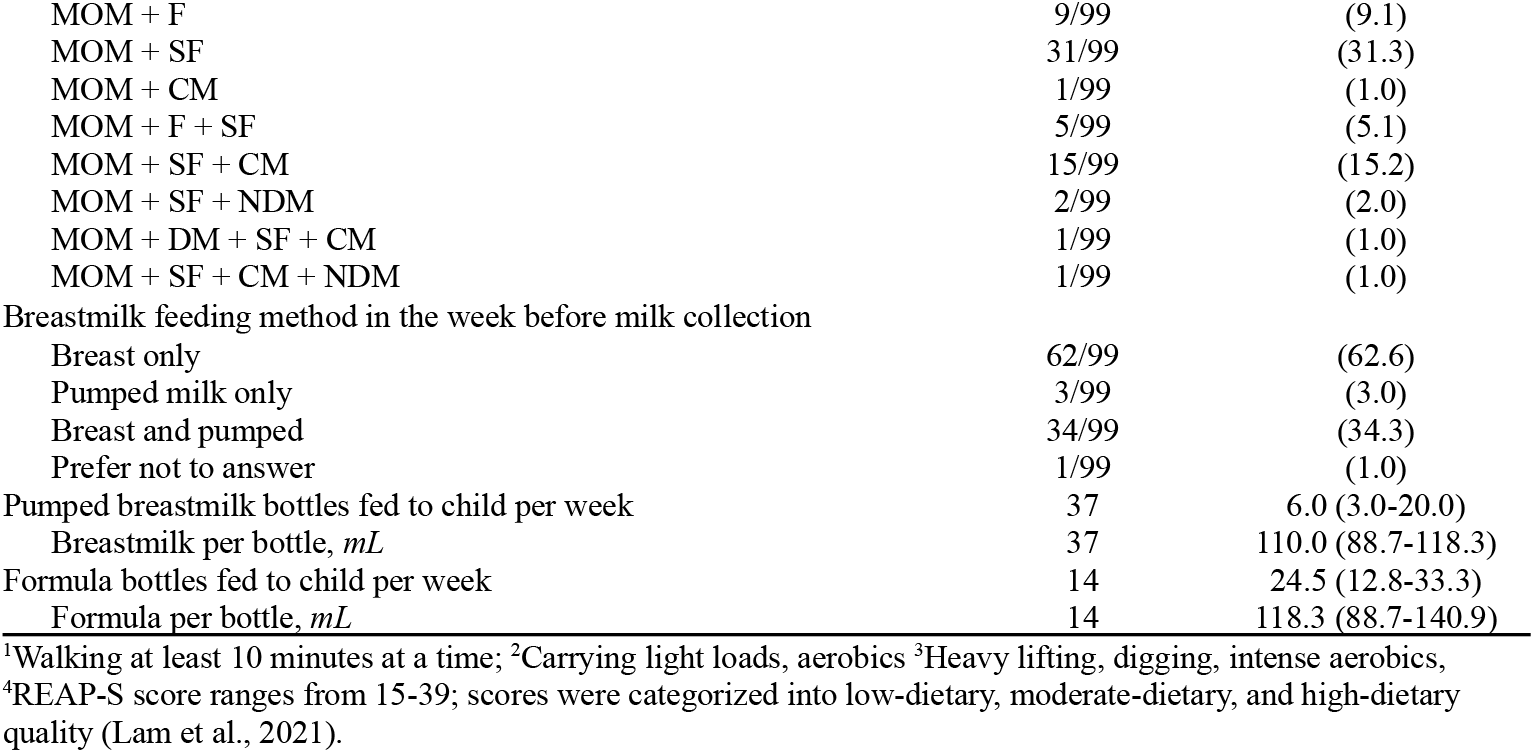
Maternal lifestyle and feeding practices in the MILC pilot study population (N=100).

|  | MILC pilot study population<br>(N=100 mothers, N=102 children) |  |
| --- | --- | --- |
|  | n, n/N | Median (IQR), or (%) |
| **Maternal lifestyle** |  |  |
| Physical activity in the LAST 7 DAYS |  |  |
| Time sitting per day, *hrs* | 83 | 35.0 (28.0-49.0) |
| Mother goes on a walk<sup>1</sup> weekly | 96/99 | (96.9) |
| Days walking<sup>1</sup> per week | 96 | 6.0 (4.0-7.0) |
| Mother does moderate<sup>2</sup> or vigorous activity<sup>3</sup> weekly | 64/94 | (68.1) |
| Days doing moderate<sup>2</sup> or vigorous<sup>3</sup> activity per week | 64 | 4.0 (2.0-5.0) |
| Average sleeping time, *hrs/day* | 99 | 7.0 (6.0-8.0) |
| Rapid Eating Assessment (REAP-S) |  |  |
| Low quality diet (score 15-19)<sup>4</sup> | 1/97 | (48.5) |
| Moderate quality diet (score 20-29)<sup>4</sup> | 29/97 | (29.9) |
| High quality diet (score 30-39)<sup>4</sup> | 67/97 | (69.1) |
| Substance use |  |  |
| Consumes alcohol (at least monthly) | 70/99 | (70.7) |
| Waits to breastfeed or pump milk after consuming alcohol | 34/70 | (48.6) |
| Time waited, *hrs* | 34 | 2.0 (1.0-2.8) |
| Consumes cannabis products (at least monthly) | 10/99 | (10.1) |
| Waits to breastfeed or pump milk after consuming cannabis | 5/10 | (50.0) |
| Time waited, *hrs* | 5 | 3.0 (3.0-6.0) |
| Consumes nicotine products (at least monthly) | 3/100 | (3.0) |
| **Feeding practices** |  |  |
| Mother previously breastfed other children | 45/100 | (45.0) |
| Mother feels they are meeting breastfeeding goals | 89/99 | (89.9) |
| Child started eating solid food | 55/99 | (55.6) |
| Age child started eating solid food, *mo* | 55 | 6.0 (5.6-6.0) |
| Child started drinking cows milk | 18/99 | (18.2) |
| Age child started drinking cows milk, *mo* | 18 | 12.2 (12.0-13.8) |
| Child started drinking non-dairy milk | 4/99 | (4.0) |
| Age child started drinking non-dairy milk, *mo* | 4 | 12.1 (10.5-13.6) |
| Child received formula in hospital | 26/99 | (26.3) |
| Child's diet in the week before milk collection |  |  |
| Mother's own breast milk (MOM) | 99/99 | (100.0) |
| Donor milk (DM) | 1/99 | (1.0) |
| Formula (F) | 14/99 | (14.1) |
| Non-dairy milk (NDM) | 4/99 | (4.0) |
| Cow's milk (CM) | 18/99 | (18.2) |
| Solid foods (SF) | 55/99 | (55.6) |
| Child's diet in the week before milk collection |  |  |
| MOM | 34/99 | (34.3) |
| MOM + F | 9/99 | (9.1) |
| MOM + SF | 31/99 | (31.3) |
| MOM + CM | 1/99 | (1.0) |
| MOM + F + SF | 5/99 | (5.1) |
| MOM + SF + CM | 15/99 | (15.2) |
| MOM + SF + NDM | 2/99 | (2.0) |
| MOM + DM + SF + CM | 1/99 | (1.0) |
| MOM + SF + CM + NDM | 1/99 | (1.0) |
| Breastmilk feeding method in the week before milk collection |  |  |
| Breast only | 62/99 | (62.6) |
| Pumped milk only | 3/99 | (3.0) |
| Breast and pumped | 34/99 | (34.3) |
| Prefer not to answer | 1/99 | (1.0) |
| Pumped breastmilk bottles fed to child per week | 37 | 6.0 (3.0-20.0) |
| Breastmilk per bottle, *mL* | 37 | 110.0 (88.7-118.3) |
| Formula bottles fed to child per week | 14 | 24.5 (12.8-33.3) |
| Formula per bottle, *mL* | 14 | 118.3 (88.7-140.9) |

<sup>1</sup>Walking at least 10 minutes at a time; <sup>2</sup>Carrying light loads, aerobics <sup>3</sup>Heavy lifting, digging, intense aerobics, <sup>4</sup>REAP-S score ranges from 15-39; scores were categorized into low-dietary, moderate-dietary, and high-dietary quality (Lam et al., 2021).

#### Participants

The median age of pilot mothers was 34 years (IQR 31-36.2). The majority were Canadian-born (85.7%), had completed a post-secondary education program (93.9%), and were of European descent (80.0%), although 55.1% reported multiple ethnicities. About half (54.1%) reported a household income over $100,000 at time of recruitment despite the majority (63.3%) of mothers being on parental leave from work. Only 2.0% of mothers reported being stay-at-home parents; the most common employment fields were healthcare (29.0%), administrative/management roles (21.0%), and education (14.0%). Nearly all mothers (96.9%) walked for at least 10 minutes a day, and the majority (68.1%) did at least one day of moderate or vigorous activity in the week before HM collection. The majority (69.1%) of mothers had a rapid eating assessment score (REAP-S) >30, which is considered a high quality diet (24). About half (50.5%) reported at least one health condition, with the most common being depression (16.2%), migraine headaches (15.2%), and irritable bowel syndrome (6.1%).) (**Tables 2 and 3**).

Children ranged from 0.3 to 36 months old at the time of enrolment (median 6.2, IQR 3.1-14.4). About half (52.9%) were male and most were born full term (37-42 weeks) (97.1%), birthed vaginally (77.5%), and healthy (80% reporting no medical conditions) at time of enrollment. Over half (60.2%) were not first born, though only 45.0% of mothers reported previously breastfeeding. All children were still consuming their mother’s HM, with 62.6% receiving HM directly at the breast only, 34.3% receiving HM both at the breast and from a bottle, and 3.0% receiving pumped and bottled HM only. 55.6% of children had started eating solid food and 14.1% consumed formula in the week before HM collection.

#### HM Samples

HM samples were collected from all mothers via electronic pump a median of 2.0 (IQR 1.0-3.3) hours after feeding their child. HM samples were processed and frozen at -80°C a median of 1.0 (IQR 0.8-1.2) hours after collection. HM macronutrient levels were consistent with similar-lactation-stage HM macronutrient levels found in literature (25,26): median protein 0.7 g/100mL (IQR 0.6-0.9); fat 3.6 g/100mL (IQR 2.7-5.2); carbohydrate 6.9 g/100mL (IQR 6.7-7.0); total energy 63.3 kcal/100mL (IQR 54.5-77.9); **Figure 2**)

**Figure 2.**
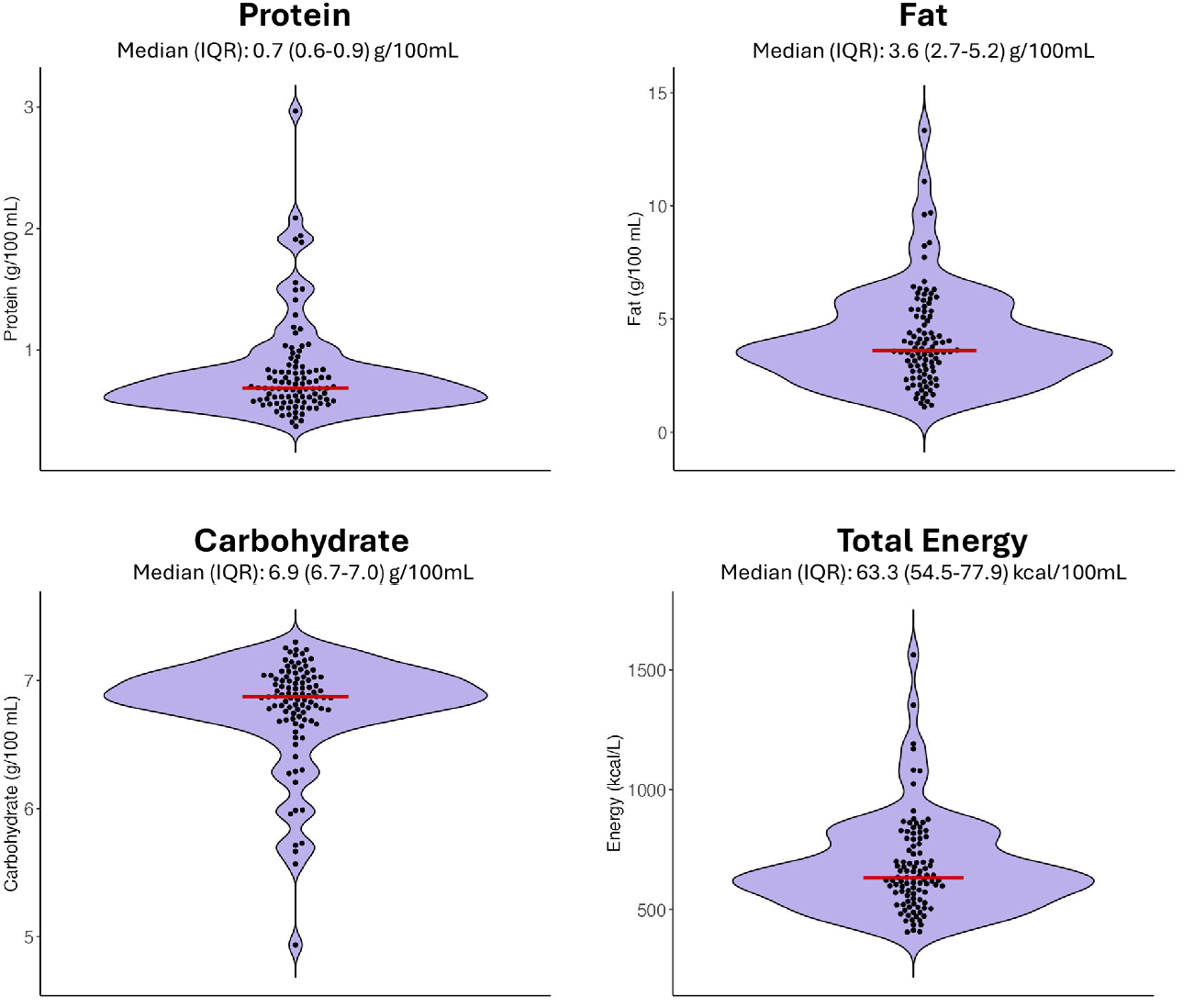
Protein, fat, carbohydrate, and total energy levels measured in human milk samples collected from MILC pilot participants (N=100) using near-infrared spectroscopy. Red bar is the median value.

#### Administrative Data Linkage

To demonstrate feasibility, provincial administrative data was linked to MILC questionnaire and HM sample data for the first n=66 recruited mothers. Within this linked sub-cohort, all were successfully linked and 63.6% (n=42/66) had lifelong administrative data available, with remaining participants having fewer years of data because they were not born in Manitoba (median 4.5 years, IQR:4.0-9.8). Infants were not linked in the pilot study because they had just been recruited and therefore no meaningful follow-up administrative data was available at the time.

### Participant and Public Involvement

The MILC study design, infrastructure, questionnaires and research foci were developed in collaboration with researchers, trainees, healthcare providers (e.g. physicians, nurses, lactation consultants), and community partners (e.g. Youville Community Health Centre) attending recurring monthly MILC Club meetings. These interdisciplinary group meetings began in 2020 to share MILC initiatives, member updates, and to catalyze knowledge mobilization. Anyone interested in MILC research can join MILC Club by contacting the MILC Club coordinator.

MILC pilot participants were asked to provide the MILC research team with open-ended feedback about their experience in the study. Feedback regarding questionnaire wording, content and structure was incorporated in subsequent revisions of the study design and questionnaires.

## DISSEMINATION AND ETHICS

MILC research results will be disseminated in peer-reviewed journal publications and at academic and clinical conferences. MILC research updates are shared via the MILC website (https://www.milcresearch.com) and through traditional and social media channels as appropriate. The MILC Club (mentioned above) is a key avenue for dissemination of research results. Through monthly meetings and an email list, members are updated about research results and collaborate to develop knowledge mobilization strategies. These include public knowledge sharing events (e.g. National Breastfeeding Week information booths and virtual “ask me anything” events), educational programming and workshops, advocacy initiatives, and community programs (e.g. Milk Mentors and the Pump Loan Library at Youville Community Health Centre). MILC Club members also engage routinely with agenda-setting organizations (e.g. International Society for Research in Human Milk and Lactation, Breastfeeding Committee for Canada, North American Board for Breastfeeding and Lactation Medicine) that are committed to translating research knowledge into policy and practice.

The “Manitoba Interdisciplinary Lactation Center (MILC): A bench-to-population human milk biorepository and research platform” study was approved by the University of Manitoba Human Research Ethics Board (RITHIM Harmony #3583, University of Manitoba Human Research Ethics Board No. HS23329 (H2019:418)) and the Provincial Health Research Privacy Committee (P2023-82). Participation in the study is voluntary and based on informed consent - participants will be able to withdraw their participation at any time.

## STRENGTHS AND LIMITATIONS

MILC fills several key gaps in HM and lactation research, which is often limited by a lack of confounding variable data (e.g. sociodemographic and maternal health factors), imprecise infant feeding data (e.g. without distinguishing mode and/or exclusivity of HM feeding), reliance on self-report and short-term followup, and a lack of HM samples to interrogate biological mechanisms. MILC is a globally unique “bench to population” facility that addresses these limitations by linking detailed questionnaires capturing health and demographic characteristics, infant feeding practices, HM samples, and lifelong provincial administrative data. Our pilot recruitment demonstrates the feasibility of MILC in collecting, analyzing, and storing HM samples, capturing unique and high resolution questionnaire data, and linking provincial administrative data. MILC is accessible to researchers worldwide and can be tailored or customized to include questionnaires or HM analyses that suit collaborator research projects. MILC also utilizes feedback from MILC Club, an interdisciplinary group of researchers, trainees, healthcare providers and community partners, as well as MILC study participants to choose meaningful research priorities and improve study design and/or processes. A MILC parent advisory group is being created to further engage research participants in shaping MILC research priorities and questionnaires.

MILC also has limitations. Some aspects of the self-report questionnaires are retrospective and subject to recall bias (e.g. recalling breastfeeding experiences with older children). The pilot study population is predominantly (80.0%) of European descent with relatively high socioeconomic status (93.9% post-secondary educated, 54.1% households generating >$100 000), compared to the general Canadian population (27), which may limit the generalizability of study results and relevance to minority populations. Multiple strategies will be employed going forward to recruit a more diverse demographic of MILC participants. Finally, the long-term maintenance and support of MILC facilities, infrastructure, and research requires long-term financial support.

## Data Availability

Data produced in this present study (i.e. MILC samples and/or questionnaire data) can be accessed with MILC Executive Committee and institutional research ethics board approval, as well as completed material and data transfer agreements.
Access to provincial administrative data requires further approval by the Manitoba Provincial Health Research Privacy Committee and institutional research ethics boards, as well as material and data transfer agreements with the Manitoba Center for Health Policy.
Access inquiries can be directed to.

## ACKNOWLEDGEMENTS AND COMPETING INTERESTS

We thank all MILC Club members (researchers, trainees, healthcare providers, and community partners) for assisting in the creation and evolution of MILC; Zahra Nouri and Shayna Geisbrecht for managing the MILC biorepository; Boluwatife Akindele for contributing to analysis; Narges Khodabandehloo and Karinne Muniz for co-ordinating the MILC Club; and Michelle Olivson for program management. We also thank the MILC study participants, without whom this research would not be possible.

MBA has received speaking honoraria from non-profit organizations that support breastfeeding (Institute for the Advancement of Breastfeeding & Lactation Education, UK Baby Friendly), and companies that produce human milk-related products (Prolacta Biosciences, Medela). She is a scientific advisor to TinyHealth and PumpKin Baby Inc. No other authors report relationships or activities that could appear to have influenced the submitted work.

## Authors Contributions

LCL, SRA, and MBA designed the study and drafted the manuscript. AJ and KF made substantial contributions to the conception and design of the biorepository and research platform. AJ recruited study participants. KF designed biorepository protocols. SRA completed statistical analysis. All authors reviewed, revised, and approved the manuscript.

